# A statewide analysis of SARS-CoV-2 transmission in New York

**DOI:** 10.1101/2021.02.20.21251598

**Authors:** Erica Lasek-Nesselquist, Navjot Singh, Alexis Russell, Daryl Lamson, John Kelly, Jonathan Plitnick, Ryan D. Pfeiffer, Nathan Tucker, Erasmus Schneider, Kirsten St. George

## Abstract

New York State, in particular the New York City metropolitan area, was the early epicenter of the SARS-CoV-2 pandemic in the United States. Similar to initial pandemic dynamics in many metropolitan areas, multiple introductions from various locations appear to have contributed to the swell of positive cases. However, representation and analysis of samples from New York regions outside the greater New York City area were lacking, as were SARS-CoV-2 genomes from the earliest cases associated with the Westchester County outbreak, which represents the first outbreak recorded in New York State. The Wadsworth Center, the public health laboratory of New York State, sought to characterize the transmission dynamics of SARS-CoV-2 across the entire state of New York from March to September with the addition of over 600 genomes from under-sampled and previously unsampled New York counties and to more fully understand the breadth of the initial outbreak in Westchester County. Additional sequencing confirmed the dominance of B.1 and descendant lineages (collectively referred to as B.1.X) in New York State. Community structure, phylogenetic, and phylogeographic analyses suggested that the Westchester outbreak was associated with continued transmission of the virus throughout the state, even after travel restrictions and the on-pause measures of March, contributing to a substantial proportion of the B.1 transmission clusters as of September 30^th^, 2020.

## Introduction

The first case of SARS-CoV-2 community transmission in New York State (and the second case overall) was documented on March 2, 2020 in the Westchester County town of New Rochelle, approximately 25 miles North of New York City. Within days, a cluster of over 90 linked cases emerged as family members, friends, and neighbors of the index case (Patient Zero) as well as healthcare workers that treated the patient tested positive (Goldstein & Salcedo 2020). From March 16^th^-July 22^nd^, the total number of cases in New York exceeded any other state (data.cdc.gov) despite voluntary quarantine measures enacted shortly after the index case tested positive, bans on large gatherings (March 12^th^; Cuomo 2020a), restaurant, bar (March 16^th^; Cuomo 2020b), and school (March 18^th^; Cuomo 2020c) closures, and the shuttering of all non-essential businesses (March 22^nd^; Cuomo 2020d). However, daily case counts dropped dramatically by late April and from early June through November, New York sustained a percent positivity rate that hovered around 1% (despite increased testing). This was largely attributable to March 22^nd^ on-pause measures and widespread adherence to face-mask and social distancing policies although the positivity rate has risen subsequently to the writing of this manuscript.

Previous genomic epidemiological investigations revealed that the majority of infections in the greater New York City metropolitan area (which includes the five boroughs of the city, and Westchester, Nassau, and Suffolk counties) derived from the Pangolin B.1 lineage prevalent in Europe and that the virus was introduced multiple times from other states and countries (Gonzalez-Reiche et al. 2020; Maurano et al. 2020). Here, we attempt to clarify the SARS-CoV-2 transmission dynamics across New York State with additional sequencing of SARS-CoV-2 samples from almost every county of New York. We identified regional variation and a shift over time in SARS-CoV-2 community composition with the Westchester outbreak contributing to the dominance of the B.1 lineage. The data suggest that after the initial surge of cases in early spring, the strict on-pause measures implemented were largely effective at curbing widespread transmission into early Autumn.

## Methods

### RNA extraction

Respiratory swabs in viral transport medium (VTM, UTM or MTM) were received at the Virology Laboratory, Wadsworth Center, for SARS-CoV-2 diagnostic testing. Total nucleic acid was extracted using either easyMAG or eMAG (bioMerieux, Durham, NC), MagNA Pure 96 and Viral NA Large Volume Kit (Roche, Indianapolis, IN), or EZ1 with the DSP Virus Kit (QIAGEN, Hilden, Germany) following manufacturers’ recommendations. Extracted nucleic acid was tested for SARS-CoV-2 RNA using the 2019-Novel Coronavirus Real-Time RT-PCR Diagnostic Panel (Centers for Disease Control and Prevention) according to the Instructions for Use on an ABI 7500Dx.

### Ion Torrent Ampliseq Panel sequencing

Viral RNA was converted to cDNA using the SuperScript VILO cDNA synthesis Kit (Thermofisher Scientific), according to the manufacturer’s instruction. Libraries were prepared on the Ion Chef System as described in the Ion AmpliSeq™ Library Preparation on the Ion Chef™ System User Guide. The samples were amplified for 17 cycles with a 4-minute extension time, using the Ion AmpliSeq SARS-CoV-2 Research Panel (Reference). Samples were then sequenced on the Ion S5 XL System (Thermofisher Scientific) using Ion 530 chips (Thermofisher Scientific) with 16 or 32 barcoded samples loaded on each chip.

### Illumina library preparation and sequencing

Whole genome amplicon sequencing of SARS-CoV-2 was performed using a modified ARTIC protocol (https://artic.network/ncov-2019). Briefly, cDNA was synthesized by combining 5 µl of RNA with SuperScript™ IV reverse transcriptase (Invitrogen, Carlsbad, CA, USA), random hexamers, dNTPs, and RNAse inhibitor, followed by incubation at 25°C for 5 minutes, 42°C for 50 minutes and 70°C for 10 minutes on a SimpliAmp thermal cycler (Thermo Fisher Scientific, Waltham, MA, USA). Two separate amplicon pools were generated by multiplex PCR with two separate premixed ARTIC V3 primer pools (Integrated DNA Technologies, Coralville, IA, USA). Additional primers to supplement those that showed poor amplification efficiency (https://github.com/artic-network/artic-ncov2019/tree/master/primer_schemes/nCoV-2019) were added separately to the pooled stocks. PCR conditions were 98°C for 30 seconds, followed by 24 cycles of 98°C for 15 seconds and 63°C for 5 minutes with a final 65°C extension for 5 minutes. Amplicons from pool 1 and pool 2 reactions were combined and purified by Ampure XP beads (Beckman Coulter, Brea, CA, USA) with a 1X bead to sample ratio and eluted in 10 mM Tris-HCl (pH 8.0). The amplicons were quantified using Quant-IT™ dsDNA Assay Kit on an ARVO™ X3 Multimode Plate Reader (Perkin Elmer, Waltham, MA, USA). Illumina sequencing libraries were generated using the Nextera DNA Flex Library Prep Kit with Illumina Index Adaptors and sequenced on a MiSeq instrument (Illumina, San Diego, CA, USA).

### Bioinformatics processing

Illumina libraries were processed with ARTIC nextflow pipeline (https://github.com/connor-lab/ncov2019-artic-nf/tree/illumina, last updated April 2020). Briefly, reads were trimmed with TrimGalore (https://github.com/FelixKrueger/TrimGalore) and aligned to the reference assembly MN908947.3 (strain Wuhan-Hu-1) by BWA (Li & Durbin 2010). Primers were trimmed with iVar (Grubaugh et al. 2018) and variants were called with samtools mpileup function (Li et al. 2009), the output of which was used by iVar to generate consensus sequences. Positions were required to be covered by a minimum depth of 50 reads and variants were required to be present at a frequency ≥ 0.75.

Ion Torrent reads were processed by the Ion AmpliSeq™ SARS-CoV-2 Research Panel with Ion SARS-CoV-2 Research Plug in package IRMAreport / Geneious 9.1.8.

### Phylogenetic and community structure analyses

We aligned our SARS-CoV-2 genomes sequenced at the Wadsworth Center and 1179 New York sequences deposited in the Global Initiative on Sharing Influenza Data (GISAID, https://gisaid.org/,) database with MAFFT v.7.450 (Katoh & Standley 2013). To qualify for inclusion in the analysis, genomes were required to have associated county metadata and to be >98% complete with fewer than 5% ambiguities. IQ-TREE v. 1.6.12 (Nguyen et al. 2015) generated a maximum likelihood phylogeny under an HKY+G4 nucleotide substitution model with 1000 ultrafast bootstrap replicates after trimming the 5’ and 3’ ends of genomes. All sequences were assigned to lineages by Pangolin v.2.0.4 (Rambaut et al. 2020). Variants were annotated by the Coronapp web tool (http://giorgilab.dyndns.org/coronapp/) (Mercatelli et al. 2020) and clade defining SNPs were identified by a custom Python script.

Potential transmission clusters were detected by TreeCluster (https://github.com/niemasd/TreeCluster; downloaded May 2020) (Balaban et al. 2019) under the threshold-free setting and an optimized branch length threshold of 0.00068. TreeCluster attempts to incorporate evolutionary distances and tree topology into defining transmission clusters with the threshold-free approach identifying a distance that maximizes the number of clusters with more than one member (Balaban et al. 2019). We tested other TreeCluster algorithms, but the threshold-free approach was the most conservative – yielding more clusters with fewer individuals (i.e. less diversity within a cluster). Although this approach showed evidence of breaking known larger transmission chains into multiple clusters (such as cases associated with the initial spread of SARS-CoV-2 in Westchester County), we chose to be conservative because we lacked associated epidemiological information for most of our samples. As a comparison, we also included the results of the single-linkage min-cut partitioning strategy, which finds the closest leaf outside a subtree and only cuts the pair of nodes if their distance does not exceed a user-specified threshold (0.00068) (Balaban et al. 2019). This algorithm led to larger clusters with more diversity and fewer sequences that lacked a cluster association, capturing deeper relationships within the tree as opposed to the tips. We also chose to include smaller transmission chains (fewer than five individuals) because they often showed inter-regional membership. Trees and annotations were visualized in FigTree v.1.4.4 (https://github.com/rambaut/figtree/) or in R v.4.0.0 (http://www.R-project.org) with the ggtree package (Yu et al. 2017).

Llama v.0.1 (https://cov-lineages.org/llama.html) placed all New York sequences generated by the Wadsworth Center within a global phylogeny of >70,000 SARS-CoV-2 genomes generated by Dr. Rob Lanfear (downloaded on 7/27/2020, https://github.com/roblanf/sarscov2phylo/). Llama incorporates query sequences into the global phylogeny and returns a subtree of closely related sequences. These contextual sequences assisted us in determining whether transmission clusters were seeded locally or via non-New York sources and were employed downstream in phylogeographic analyses.

The R package Picante (Kembel et al. 2010) calculated alpha-diversity metrics (species richness and divergence) for the ten regions of New York State (Capital District, Central New York, Finger Lakes, Long Island, Mid-Hudson, Mohawk Valley, New York City, North Country, Southern Tier, and Western New York). The mean pairwise distance (mpd) and mean nearest taxon distance (mntd) functions determined the presence of phylogenetic structuring within each region (more or less divergence than expected by chance). Both functions calculate average distances among taxa within a community (all pairwise distances or the shortest distances) and compare the expected amount of phylogenetic relatedness against a null distribution (Webb et al. 2008) generated from 1000 community randomizations. Mpd is believed to capture community structure more deeply within the tree while the mntd identifies structure closer to the tips (Webb et al. 2008; Tucker et al. 2017). Calculations were performed at the Pangolin lineage level and for all threshold-free transmission clusters represented in the New York tree.

We employed weighted (Lozupone et al. 2007) and unweighted UniFrac tests (Lozupone & Knight 2005) in mothur v.1.44.3 (Schloss et al. 2009) to evaluate beta-diversity (or the dissimilarity of viral communities) between regions. These methods rely on a phylogenetic tree to measure evolutionary divergence between communities with the weighted method incorporating abundance information as well. Weighted and unweighted UniFrac tests can be employed to understand different aspects driving community structure (Lozupone et al. 2007). The weighted UniFrac test detects shifts in relative abundances between communities while the unweighted test is suited to identifying founding populations (Lozupone et al. 2007). Whether regions display significantly different viral communities is evaluated by randomizing the communities on the tree and recalculating UniFrac distances to determine the proportion of trees with a distance greater than or equal to the observed data. P-values were adjusted for multiple testing by the false discovery rate (FDR) with the p.adjust function in R.

### Phylogeographic analyses

Phylogeographic analyses were conducted in Nextstrain v. 2.0.0.post1 (Hadfield et al. 2018) using custom builds and the Nextstrain pipeline. The builds included New York SARS-CoV-2 genomes sequenced by the Wadsworth Center and a subset of New York sequences obtained from GISAID. To place New York sequences in a global context, Nextstrain sub-sampled SARS-CoV-2 genomes from the rest of the world based on genomic proximity, geography, and temporal distribution, producing phylogenies of roughly 2700-3700 genomes. Sequences were required to be ≥ 29 Kb in length with ≤ 3% ambiguities and were masked at the first 100 and last 50 positions. Additionally, problematic positions 13402, 24380, and 24390 were masked before aligning with MAFFT v.7.470. Maximum likelihood phylogenetic reconstruction was performed in IQ-TREE v.2.0.3 with ancestral traits and divergence times inferred by TreeTime v.0.7.6 (Sagulenko et al. 2018; Hadfield et al. 2018) and strain Wuhan-Hu-1/2019 (MN908947.3 or GISAID number EPI_ISL_402125) set as the root. Trees spanned from December 2019 to June 2020 and were visualized in Auspice.

Three phylogeographic analyses were also conducted in Beast v.2.6.2 9 (Bouckaert et al. 2019; Lemey et al. 2009) under an exponential population model and geographic locations (America/USA, Europe, Asia, Oceania, and New York) set as discrete traits to predict the source of New York transmission clusters (as defined by the threshold-free approach in TreeCluster) and to elucidate the breadth of the Westchester outbreak. Included in the first two analyses were the earliest sequences from the Westchester outbreak (from individuals known to be directly associated with the index), representatives from a subset of New York B.1.X transmission clusters (as defined by the threshold-free approach in TreeCluster), a subset of “contextual” sequences obtained from Nextstrain builds (including the earliest B.1 genomes identified in Europe and the United States), and genomes identified by Llama as putatively derived from out-of-state transmission. Two independent runs were performed with representatives from 116/303 or 94/303 B.1.X transmission clusters that spanned the diversity of B.1.X lineages in New York. The third analysis included the earliest representatives from each B.1.X threshold-free transmission cluster that appeared between March and July (n=259, excluding some that were consistently identified as out-of-state introductions by the first two analyses), the index genome and those with an epidemiological link, and a subset of genomes from the United States and Europe. Non-New York B.1.X genomes were randomly selected between February to July from GISAID, with the number of genomes sampled each month approximately equal to those of New York for a total of 596 sequences (more genomes were randomly sampled from the United States as reduced transmission between countries after nationwide lockdowns made international importations less likely (Magalis et al. 2020)). All analyses were run under an HKY nucleotide substitution model with a gamma distribution and four rate categories. The clock rate was fixed at 8×10^−04^ substitutions per site per year (Nie et al. 2020; Rambaut, Andrew 2020) under a normal prior with standard deviation set to 1×10^−05^. Runs were monitored for convergence in Tracer v.1.7.1 (Rambaut et al. 2018) and stopped when effective sample sizes were ≥ 200 for all parameters. Maximum clade credibility trees were generated by TreeAnnotator v.2.6.2 (Bouckaert et al. 2019; Heled & Bouckaert 2013) with a 10% burn-in and node heights equal to the common ancestor (in order to avoid negative branch lengths). BALTIC (https://github.com/evogytis/baltic) identified all New York sequences that descended from a non-New York node. Hamming distances, the number of substitutions between pairs of sequences excluding ambiguous positions and gaps, were calculated with snp-dists v. 0.7.0 (https://github.com/tseemann/snp-dists) to determine the median number of differences between genomes putatively linked to the Westchester outbreak and those independently introduced to New York State.

## Results and Discussion

### SARS-CoV-2 diversity declines across New York State as regional phylogenetic structuring emerges

Our analyses included 675 genomes sequenced by the Wadsworth Center from 56 out of 62 counties and all ten regions of New York. In total, 1854 SARS-CoV-2 genomes were analyzed, sampled from 57 New York counties between March 2^nd^ and September 30^th^, 2020 (Supplementary Table 1). The proportion of samples derived from males and females was approximately equal (48% vs. 52%) with ∼60% from those ≥ 50 years of age (Supplementary Table 1). The most heavily sampled regions were New York City, the Mid-Hudson (which includes Westchester County, the site of the initial community outbreak in New York) and Long Island – collectively referred to as “downstate” in this study (Figure 1). “Upstate” sequences – those from the North Country, Capital District, Central New York, Southern Tier, Finger Lakes, Mohawk Valley, and Western New York regions – represented about 25% of the dataset (Figure 1). Sampling was uneven among the ten regions, with downstate representing over 80% of the samples in March and April but upstate representing most samples for the remaining months (Supplementary Table 1). The number of sequences also decreased as a function of time, with 55% of the sequencing effort occurring in March and over 80% occurring in March and April combined (Supplementary Table 1).

**Figure 1.**
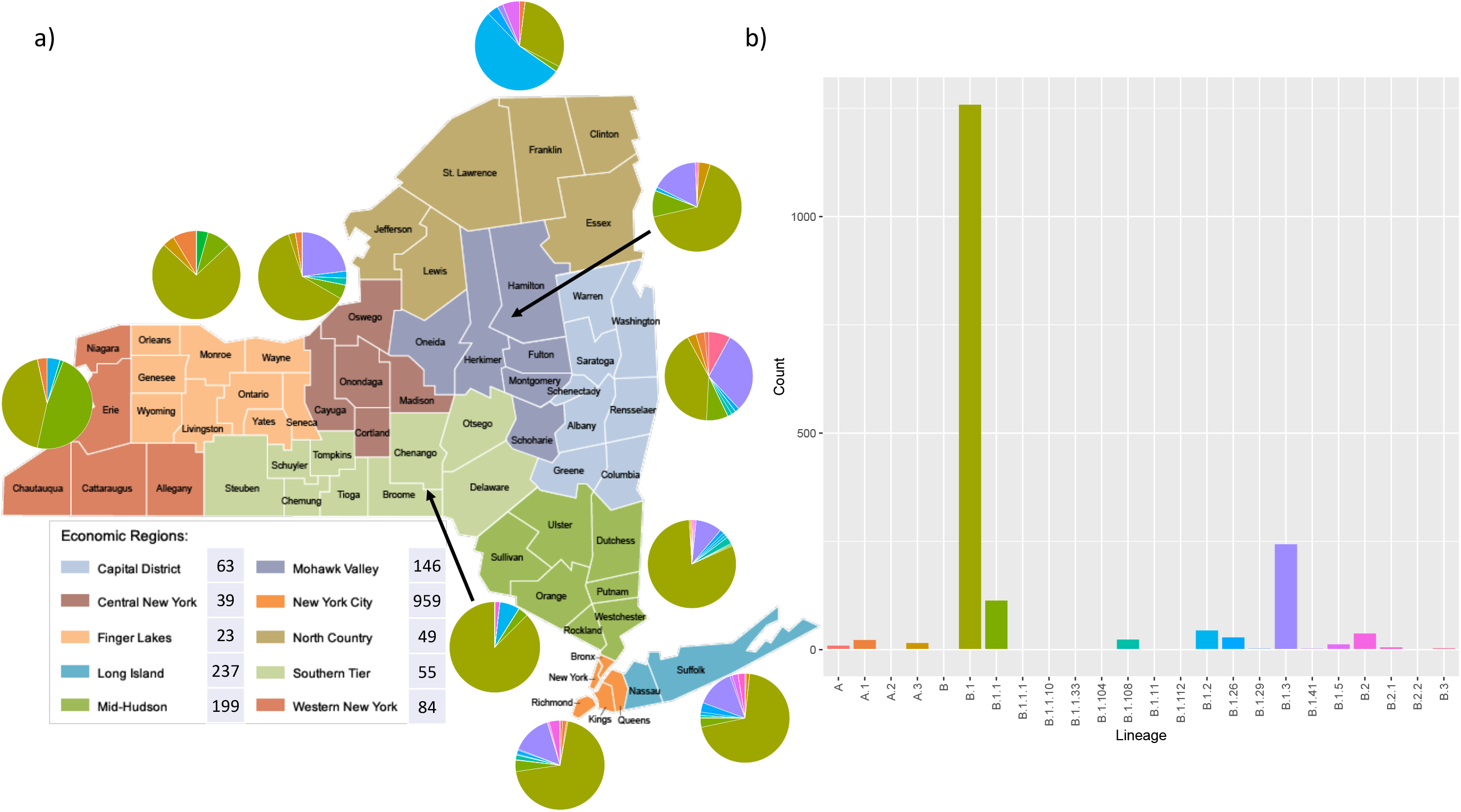
Representation of SARS-CoV-2 lineages within New York State. a) A map of the ten New York regions, the total number of SARS-CoV-2 genomes sequenced from each region, and the breakdown of lineages detected in each region (pie graphs). b) Barplot representing the number of sequences from each lineage identified in our New York State dataset of 1859 genomes. Map adapted from http://wwe1.osc.state.ny.us.

Although 24 Pangolin lineages were identified in the New York State dataset, 68% were classified as B.1 and 94% had a B.1 or descendant Pangolin lineage designation (those with a “B.1.X” nomenclature; Figure 1). The B.1 lineage was geographically and temporally ubiquitous while others displayed a more checkered appearance (Figure 2). For example, the B.1.2 lineage was significantly associated with the North Country (χ^2^test p-value < 2.2×10^−16^), which harbored more than half of the B.1.2 genomes but contributed to less than 3% of the total dataset (Supplementary Table 2). Similarly, there was a significant association between Western New York and the B.1.1 lineage (χ^2^ test, p-value < 2.2×10^−16^). SARS-CoV-2 sequences from Western New York comprised only 4.5% of the dataset (supplementary Table 2) but 30% of B.1.1 genomes derived from this region. The observed regional associations of some lineages could be due to biased and limited sampling. However, the North Country represents the most robustly sampled region in the dataset with four percent of the positive cases sequenced (Supplementary Table 2). Although four B.1.2 genomes were linked to a senior living facility, several genomes appeared to be sampled from the greater community. Less than one percent of the positive cases from Western New York were sequenced (Supplementary Table 2), but the wide distribution of sample ages suggests B.1.1 might have been the dominant lineage circulating in the general population. Thus, despite sampling limitations, some regions appear to be characterized by SARS-CoV-2 lineages that were less representative of the entire state, implicating localized viral transmission as a greater influence on SARS-CoV-2 composition than inter-regional spread.

**Figure 2.**
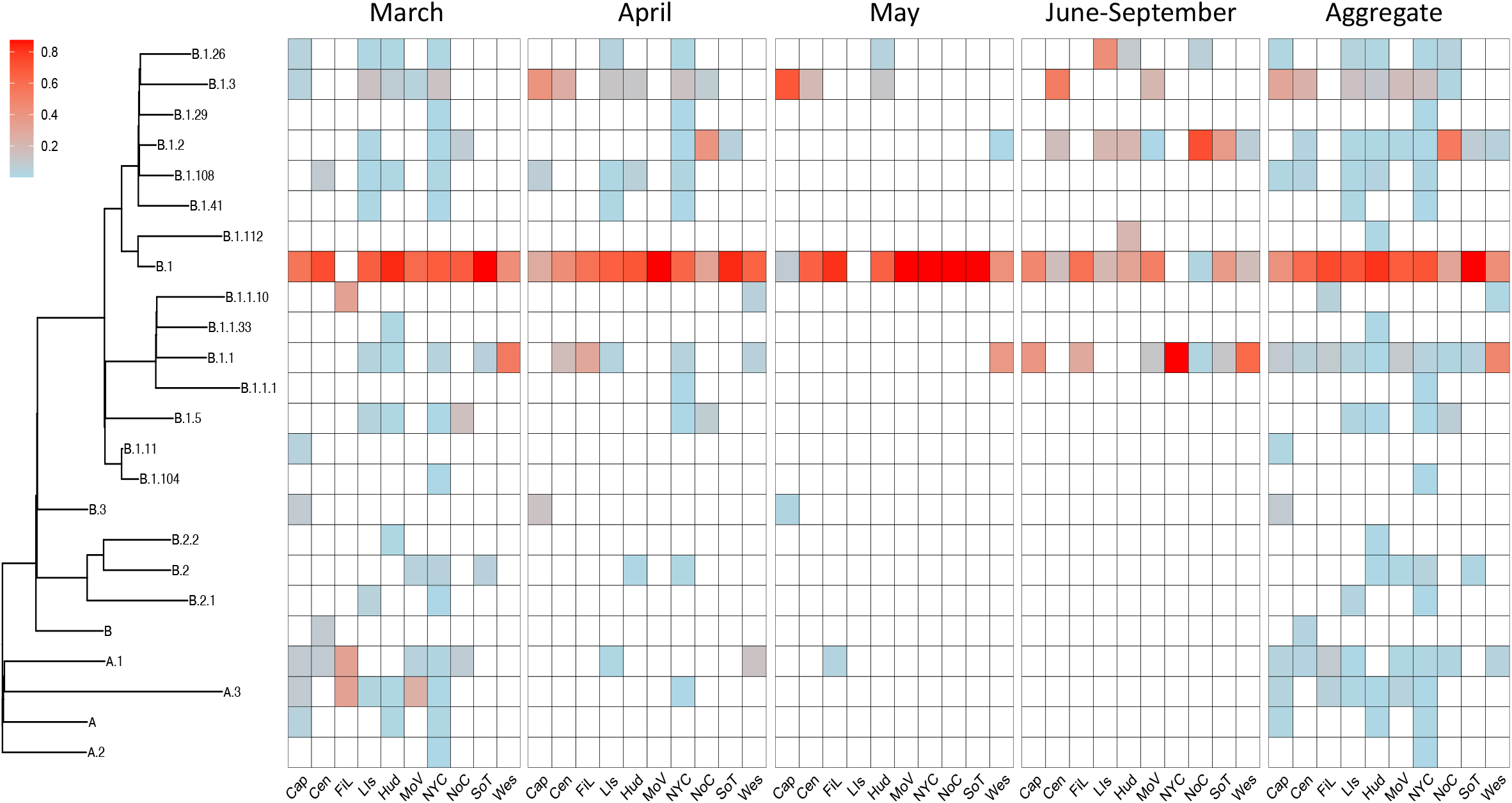
Heatmap of SARS-CoV-2 lineages in the ten regions of New York State. Heatmap of the relative frequencies of the SARS-CoV-2 lineages represented in the associated phylogeny for each New York State region from March through September 2020 and aggregated over all months. The heatmap colors indicate lineage frequencies per region per month or aggregated per region (note: 0.8 frequency represents 0.8-1.0). Tallies from June-September were combined due to smaller samples sizes. Cap, Capital District; Cen, Central; FiL, Finger Lakes; Lis, Long Island; Hud, Hudson; MoV, Mohawk Valley; NYC, New York City; NoC, North Country; SoT, Southern Tier; Wes, Western.

Overall, the diversity of observed lineages decreased with time since the beginning of the New York epidemic in March (Figure 2). This is particularly apparent by June-September, where sequence diversity is solely concentrated in the B.1.X portion of the tree (Figure 2). This does not take into account the more limited sequencing that occurred during later months, which might hinder the ability to detect variants circulating at low frequency. In fact, three regions with the greatest number of lineages in March (New York City, Mid-Hudson, and Long Island) had no data available for various later months (as of the time of writing). The number of lineages detected per month was significantly correlated with the number of samples sequenced (Spearman rank correlation coefficient r=0.74, p-value=3.9×10^−08^) and randomly down-sampling earlier months (March and April) to the size of May and June datasets revealed significant decreases in the number of lineages identified (one-sample t-test p-values < 2.2×10^−16^). However, rarefaction analyses demonstrated that while lineages continued to be identified as more regions were sampled for March and April, lineage richness remained essentially flat or unchanged for May and June-September (Supplementary Figure 1), which suggests that sampling bias did not substantially influence the underlying loss of diversity observed.

Phylogenetic structuring was detected at the Pangolin lineage level for the North Country and the Southern Tier (Table 1), where the dominance of the B.1.2 lineage in the North Country and the absence of A lineages in the Southern Tier (Figure 2) led to more similar sequences within each region than expected by chance. The lack of significant structuring for the remaining eight New York regions captures the presence of both A and B lineages within these regions (Table 1), leading to sequences distributed across the tree (Figure 2). On a monthly basis, phylogenetic structuring was not detected for any region at the beginning of the epidemic in New York State (Supplementary Table 3). However, alpha-diversity significantly declined by June for many regions (Supplementary Table 3), consistent with the lack of A and non-B.1.X lineages in later months. This potentially reflects the changing transmission dynamics that occurred within New York State once international travel restrictions and New York State on-pause measures were imposed (March 19^th^ and 22^nd^, respectively). While the initial presence of SARS-CoV-2 in New York derived from several sources (Gonzalez-Reiche et al. 2020; Maurano et al. 2020), localized transmission events appeared to be responsible for the continued presence of the virus during summer months, including September.

**Table 1.** Phylogenetic structuring results at the lineage level based on mpd and mntd values for the ten regions of New York. Ntaxa, number of lineages; observed, observed mpd or mntd value; random, the mean mpd or mntd value after 1000 randomizations of the taxa on the tree; sd, the standard deviation of the randomized means; z-score, the tendency to show more (negative values) or less (positive values) phylogenetic structuring by chance. P-values < 0.050 are considered significant.

|  | mpd |  |  |  |  |  | mntd |  |  |  |  |
| --- | --- | --- | --- | --- | --- | --- | --- | --- | --- | --- | --- |
|  | ntaxa | observed | random | sd | z score | p-value | observed | random | sd | z score | p-value |
| Capital District | 10 | 2.61E-04 | 3.18E-04 | 6.85E-05 | -0.82 | 0.21 | 2.04E-04 | 2.56E-04 | 6.50E-05 | -0.80 | 0.19 |
| Central | 7 | 1.60E-04 | 2.47E-04 | 6.51E-05 | -1.34 | 0.09 | 1.64E-04 | 2.79E-04 | 8.89E-05 | -1.29 | 0.08 |
| Finger Lakes | 5 | 2.06E-04 | 1.91E-04 | 4.85E-05 | 0.30 | 0.61 | 3.30E-04 | 3.06E-04 | 1.09E-04 | 0.22 | 0.60 |
| Long Island | 11 | 1.49E-04 | 2.13E-04 | 5.20E-05 | -1.23 | 0.11 | 1.66E-04 | 2.53E-04 | 8.83E-05 | -0.98 | 0.14 |
| Mid-Hudson | 13 | 9.04E-05 | 1.49E-04 | 3.87E-05 | -1.51 | 0.05 | 1.46E-04 | 2.46E-04 | 9.86E-05 | -1.02 | 0.12 |
| Mohawk Valley | 7 | 1.85E-04 | 2.26E-04 | 5.95E-05 | -0.69 | 0.25 | 1.84E-04 | 2.80E-04 | 9.47E-05 | -1.02 | 0.13 |
| New York City | 17 | 1.58E-04 | 2.13E-04 | 5.24E-05 | -1.06 | 0.15 | 1.63E-04 | 2.34E-04 | 8.68E-05 | -0.82 | 0.21 |
| North Country | 7 | 1.23E-04 | 2.74E-04 | 7.43E-05 | -2.03 | 0.01 | 1.54E-04 | 2.80E-04 | 8.30E-05 | -1.51 | 0.04 |
| Southern Tier | 4 | 5.56E-05 | 1.01E-04 | 2.92E-05 | -1.55 | 0.05 | 1.48E-04 | 3.32E-04 | 1.32E-04 | -1.39 | 0.05 |
| Western | 5 | 1.89E-04 | 2.57E-04 | 7.88E-05 | -0.86 | 0.22 | 1.99E-04 | 3.07E-04 | 9.78E-05 | -1.10 | 0.13 |

### The Westchester outbreak represents the largest transmission cluster in New York State, reaching numerous areas, but clusters become more regionally distinct through time

The threshold-free approach in TreeCluster identified 325 New York transmission clusters (303 of which were assigned to B.1.X lineages) from March to September with 44% of genomes belonging to a cluster of ≥ 3 members and 36% having no cluster affiliation. Genomes from the outbreak in Westchester County were assigned to the B.1 lineage and the Nextstrain 20C clade based upon sequences from the index case and those with direct affiliation to Patient Zero. The genome of the index case and those of the 20C clade deviate from the Wuhan-1 reference by six mutations: C241T, C3037T, C14408T, A23403G, C1059T, and G25563T. The first four mutations characterize the B.1 lineage while G25563T was present in the 20C parent lineage circulating in Europe by January and February 2020. The C1059T mutation is present in only one other genome collected in the United States prior to March, making It possible that most genomes with this signature mutation are derived from the initial Westchester outbreak. However, the threshold-free approach separated SARS-CoV-2 genomes from the Westchester outbreak into at least four clusters (based on the phylogenetic position of the index and epidemiologically linked genomes) that contained a total of 251 sequences, spanned 20 counties (12 upstate), and lasted from March 2 until early May 2020 (Figure3). The index genome fell within a clade of 181 identical sequences, which deviated from the genomes in the other three clusters by 1 SNP (excluding ambiguous positions led to identical sequences among the two largest clusters). The SARS-CoV-2 genome from a known epidemiologically linked case to the index was part of a separate transmission cluster and deviated from that of the index by a single SNP not detected in any other New York genome examined. This raises the possibility that there are other genomes related to the outbreak that were not included in our estimate.

The single-linkage cluster approach yielded 99 transmission clusters with 75% of sequences belonging to a cluster of three or more members and only 19% lacking a cluster affiliation. While the number of clusters with two or three members did not change substantially (Supplementary Figure 2), genomes that were not assigned to a cluster by the threshold-free approach tended to be subsumed into the Westchester outbreak. The Westchester outbreak as defined by the single-linkage algorithm and the placement of the index genome contained 877 sequences, spanned 43 counties (32 upstate) and lasted 92 days (Figure 3). There was a median Hamming distance of two between genomes of this cluster, consistent with results for linked genomes in previous analyses (Dearlove et al. 2020). This suggests that the initial community spread of SARS-CoV-2 in Westchester County was responsible for at least 13.5% to 47% of the cases represented in our dataset and 30-60% of all cases affiliated with a transmission cluster, with widespread distribution across the state. While the threshold-free approach likely underestimates the number of genomes linked to the initial Westchester outbreak by only identifying clusters at the tips of the tree instead of sustained transmission chains, the single-linkage strategy might misassign genomes to this cluster by capturing the shared ancestry of B.1.X sequences.

**Figure 3.**
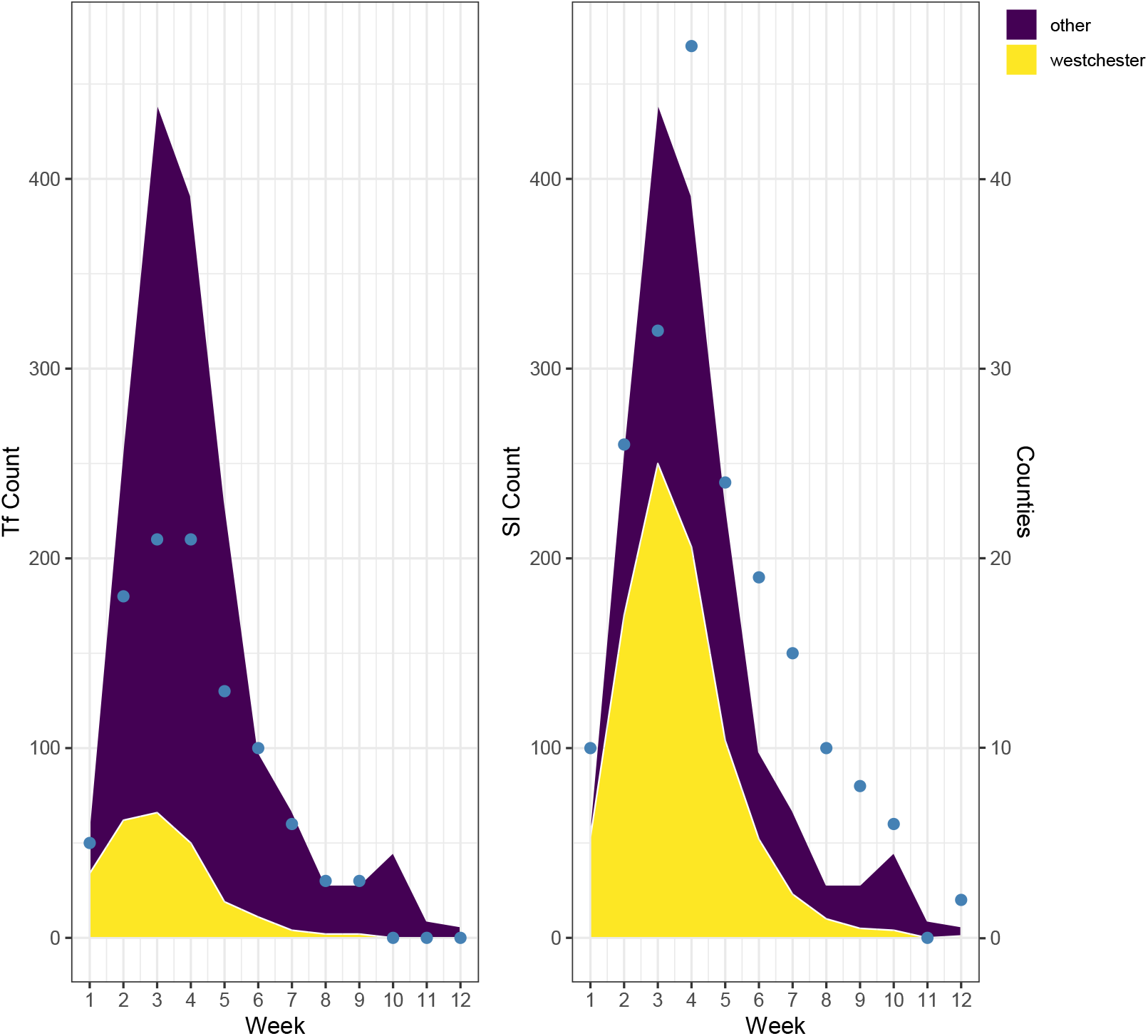
Number of genomes assigned to Westchester outbreak. Area maps of the number of genomes associated with the Westchester outbreak and all other clusters as inferred by the threshold-free (Tf) and single-linkage (Sl) clustering strategies from early-March to early-June. Blue circles indicate the proportion of counties included in the Westchester outbreak cluster for each week.

At the transmission cluster level (defined by the threshold-free approach), downstate areas of New York City, Long Island, and Mid-Hudson consistently exhibited significant phylogenetic structuring, either aggregated over all months or on a monthly basis for mntd and mpd calculations (Table 2 and Supplementary Table 4). These results appear to capture the dynamics of the Westchester outbreak as well as upstate SARS-CoV-2 transmission patterns. Sequences from upstate could be found interspersed throughout the New York State phylogeny - in predominantly downstate clades, such as the Westchester outbreak and in regionally specific clades - leading to larger distances among SARS-CoV-2 genomes from these communities (Figure 4). While outbreaks downstate (such as the one in Westchester County) contributed to seeding upstate transmission chains, the effects of upstate transmission to downstate appear minimal. The sparse sampling of New York City, Long Island, and the Mid-Hudson after May might also contribute to the pattern observed and prevents us from evaluating the full extent of SARS-CoV-2 exchange between the two areas. Sequences from upstate regions span all months considered and therefore encompass a greater proportion of the evolution that has occurred along the New York phylogeny (Figure 4). In contrast, the diversity detected downstate is largely confined to the early months of the pandemic in New York State (Figure 4).

**Table 2.** Phylogenetic structuring results at the transmission cluster level based on mpd and mntd values for the ten regions of New York. Ntaxa, number of transmission clusters; observed, observed mpd or mntd value; random, the mean mpd or mntd value after 1000 randomizations of the taxa on the tree; sd, the standard deviation of the randomized means; z-score, the tendency to show more (negative values) or less (positive values) phylogenetic structuring by chance. P-values < 0.050 are considered significant.

|  | mpd |  |  |  |  |  | mntd |  |  |  |  |
| --- | --- | --- | --- | --- | --- | --- | --- | --- | --- | --- | --- |
|  | ntaxa | observed | random | sd | z score | p-value | observed | random | sd | z score | p-value |
| Capital District | 26 | 3.90E-04 | 2.84E-04 | 5.15E-05 | 2.04 | 0.98 | 1.28E-04 | 1.27E-04 | 2.63E-05 | 0.01 | 0.54 |
| Central | 12 | 2.29E-04 | 2.61E-04 | 6.74E-05 | -0.47 | 0.35 | 1.06E-04 | 1.55E-04 | 4.53E-05 | -1.08 | 0.15 |
| Finger Lakes | 7 | 3.75E-04 | 2.56E-04 | 8.03E-05 | 1.49 | 0.91 | 1.58E-04 | 1.76E-04 | 5.81E-05 | -0.32 | 0.42 |
| Long Island | 94 | 1.74E-04 | 2.85E-04 | 4.21E-05 | -2.63 | 0.00 | 5.75E-05 | 8.85E-05 | 1.53E-05 | -2.02 | 0.00 |
| Mid-Hudson | 44 | 1.20E-04 | 2.60E-04 | 6.40E-05 | -2.18 | 0.00 | 5.05E-05 | 1.12E-04 | 3.75E-05 | -1.64 | 0.00 |
| Mohawk Valley | 32 | 4.39E-04 | 2.83E-04 | 5.13E-05 | 3.06 | 0.99 | 1.06E-04 | 1.23E-04 | 2.53E-05 | -0.65 | 0.28 |
| New York City | 214 | 1.72E-04 | 2.82E-04 | 4.74E-05 | -2.33 | 0.00 | 3.74E-05 | 6.85E-05 | 1.40E-05 | -2.22 | 0.00 |
| North Country | 15 | 3.16E-04 | 2.66E-04 | 6.58E-05 | 0.76 | 0.76 | 1.10E-04 | 1.48E-04 | 4.08E-05 | -0.93 | 0.19 |
| Southern Tier | 21 | 2.77E-04 | 2.78E-04 | 5.78E-05 | -0.01 | 0.54 | 1.50E-04 | 1.37E-04 | 3.21E-05 | 0.42 | 0.69 |
| Western | 22 | 4.80E-04 | 2.82E-04 | 5.57E-05 | 3.56 | 1.00 | 1.03E-04 | 1.33E-04 | 3.03E-05 | -1.01 | 0.15 |

**Figure 4.**
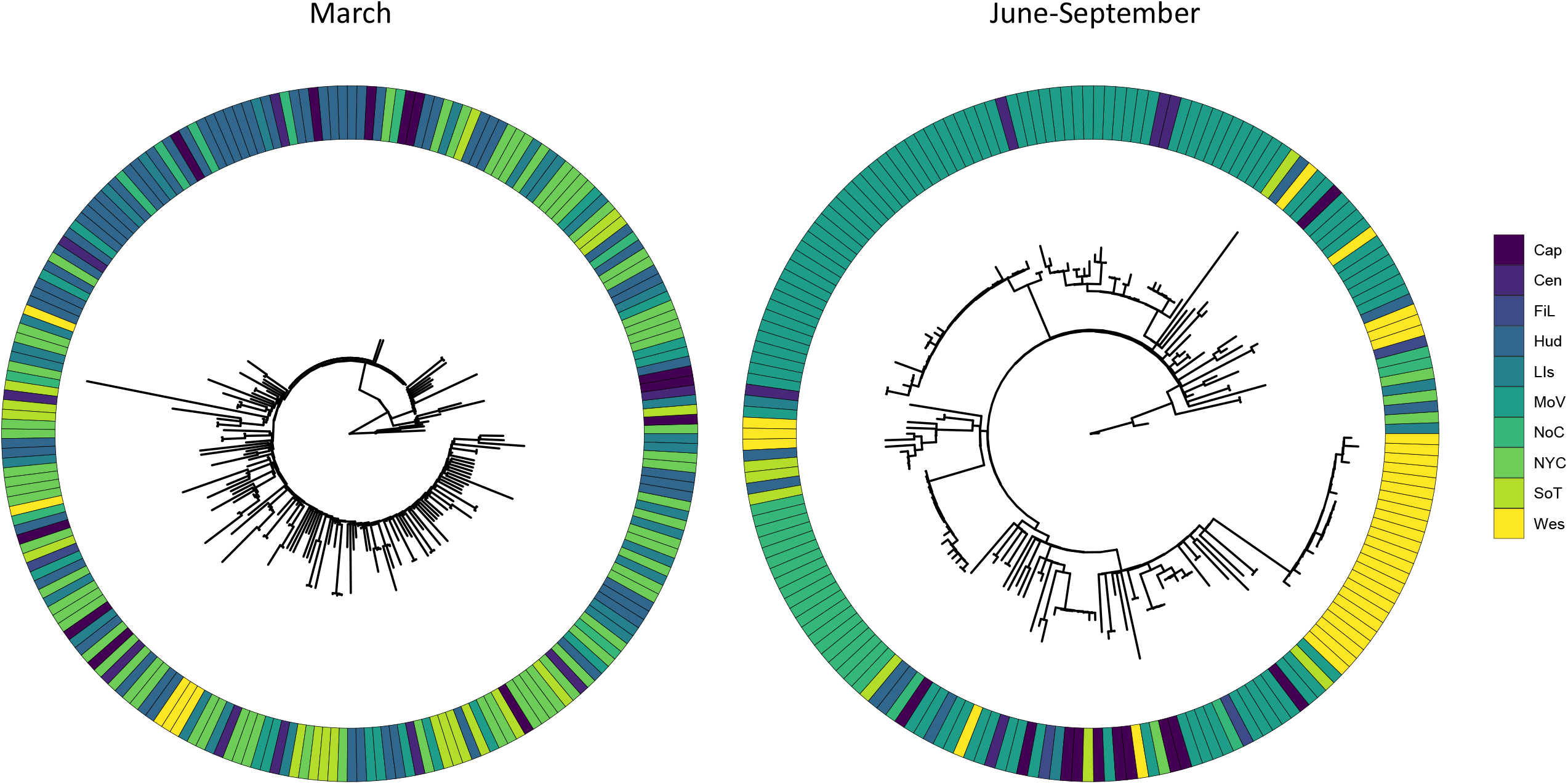
Evolutionary relationships among SARS-CoV-2 genomes sampled in New York State. Maximum likelihood phylogenies depicting relationships among New York State SARS-CoV-2 genomes on a monthly basis to highlight the increased structuring by region as the months progressed. The March dataset was randomly downsampled for ease of visualization and June-September were combined due to smaller sample sizes. Cap, Capital District; Cen, Central; FiL, Finger Lakes; Lis, Long Island; Hud, Hudson; MoV, Mohawk Valley; NYC, New York City; NoC, North Country; SoT, Southern Tier; Wes, Western.

Weighted UniFac analyses of beta-diversity indicated that all regions harbored phylogenetically distinct viral communities (Table 3). Stratifying by month uncovered a more complicated pattern, where viral communities became generally more distinct as a function of time (Figure 4 and Supplementary Table 5). This could be due to limited travel within New York and between New York and other states after March, which restricted SARS-CoV-2 transmission to more regional or even county-level scales. Results could also be influenced by differences in sampling strategies between months, with broader surveillance efforts in earlier months and an increased focus on outbreaks after May. However, transmission dynamics had also changed by May, with the number of infections declining drastically and localized outbreaks comprising a larger proportion of positive cases. For example, the percentage of transmission clusters composed of genomes from more than one region declined from 29% in March to 4% in May. By June, no clusters contained genomes from more than one region, suggesting on-pause measures were largely effective, although the New York City metropolitan area was severely under-sampled. Regardless, this trend is similar to the dynamics identified at the global level, which showed a decline in the number and size of transmission clusters as well as the number of multi-country clusters after the implementation of national lockdowns and travel restrictions (Magalis et al. 2020).

Unweighted UniFrac tests largely mirrored the results of weighted UniFrac analyses (Table 3 and Supplementary Table 5). Additionally, unweighted UniFrac results were consistent with the conclusion that downstate regions of New York City, the Mid-Hudson, and Long Island served as significant viral sources for all upstate areas except Western New York (Figure 5). This is observed most acutely for March, when SARS-CoV-2 beta-diversity between five of the six upstate regions (the Finger Lakes was excluded because of small sample size) and at least one downstate region was not significantly different than expected by chance. Further, the SARS-CoV-2 viral communities of four upstate regions did not significantly diverge from any of those downstate (Figure 5). In contrast, upstate regions showed less similarity to each other (Figure 5 and Supplementary Table 5). The SARS-CoV-2 viral community of Western New York remained consistently different from the rest of New York, suggesting that transmission from downstate and other upstate regions did not heavily influence its composition.

**Table 3.** Results from weighted and unweighted UniFrac analyses comparing beta-diversity between the ten New York regions. Scores, estimates of beta-diversity for group comparison where weighted estimates consider taxon abundance; p-values, p-values associated with 1000 randomizations of taxa labels across the New York phylogeny to determine whether communities are more dissimilar than by chance; FDR, adjusted p-value for multiple testing using the false discovery rate. Cap, Capital District; Cen, Central New York; Fil, Finger Lakes; Hud, Mid-Hudson; LIs, Long Island; MoV, Mohawk Valley; Noc, North Country; NYC, New York City; SoT, Souther Tier; Wes, Western New York.

| Regions | Weighted |  |  | Unweighted |  |  |
| --- | --- | --- | --- | --- | --- | --- |
|  | Score | p-value | FDR | Score | p-value | FDR |
| Cap-Cen | 0.60 | 0.001 | 0.00 | 0.88 | 0.00 | 0.00 |
| Cap-FiL | 0.60 | 0.012 | 0.01 | 0.83 | 0.03 | 0.03 |
| Cap-Hud | 0.70 | 0.001 | 0.00 | 0.90 | 0.01 | 0.01 |
| Cap-LIs | 0.63 | 0.001 | 0.00 | 0.89 | 0.01 | 0.01 |
| Cap-MoV | 0.65 | 0.001 | 0.00 | 0.88 | 0.00 | 0.00 |
| Cap-NoC | 0.80 | 0.001 | 0.00 | 0.88 | 0.00 | 0.00 |
| Cap-NYC | 0.59 | 0.001 | 0.00 | 0.94 | 0.08 | 0.08 |
| Cap-SoT | 0.79 | 0.001 | 0.00 | 0.91 | 0.00 | 0.00 |
| Cap-Wes | 0.81 | 0.001 | 0.00 | 0.93 | 0.00 | 0.00 |
| Cen-FiL | 0.72 | 0.001 | 0.00 | 0.89 | 0.00 | 0.00 |
| Cen-Hud | 0.60 | 0.001 | 0.00 | 0.93 | 0.04 | 0.04 |
| Cen-LIs | 0.54 | 0.03 | 0.03 | 0.91 | 0.09 | 0.10 |
| Cen-MoV | 0.63 | 0.001 | 0.00 | 0.91 | 0.00 | 0.00 |
| Cen-NoC | 0.76 | 0.001 | 0.00 | 0.86 | 0.00 | 0.00 |
| Cen-NYC | 0.52 | 0.007 | 0.01 | 0.96 | 0.05 | 0.06 |
| Cen-SoT | 0.72 | 0.001 | 0.00 | 0.92 | 0.00 | 0.00 |
| Cen-Wes | 0.80 | 0.001 | 0.00 | 0.90 | 0.00 | 0.00 |
| FiL-Hud | 0.75 | 0.001 | 0.00 | 0.94 | 0.10 | 0.11 |
| FiL-LIs | 0.69 | 0.001 | 0.00 | 0.91 | 0.35 | 0.35 |
| FiL-MoV | 0.67 | 0.001 | 0.00 | 0.90 | 0.00 | 0.00 |
| FiL-NoC | 0.80 | 0.001 | 0.00 | 0.86 | 0.00 | 0.00 |
| FiL-NYC | 0.67 | 0.001 | 0.00 | 0.97 | 0.21 | 0.21 |
| FiL-SoT | 0.75 | 0.001 | 0.00 | 0.91 | 0.00 | 0.00 |
| FiL-Wes | 0.78 | 0.001 | 0.00 | 0.91 | 0.00 | 0.00 |
| Hud-LIs | 0.47 | 0.001 | 0.00 | 0.89 | 0.00 | 0.00 |
| Hud-MoV | 0.71 | 0.001 | 0.00 | 0.92 | 0.00 | 0.00 |
| Hud-NoC | 0.73 | 0.001 | 0.00 | 0.93 | 0.00 | 0.00 |
| Hud-NYC | 0.45 | 0.001 | 0.00 | 0.92 | 0.00 | 0.00 |
| Hud-SoT | 0.64 | 0.001 | 0.00 | 0.92 | 0.00 | 0.00 |
| Hud-Wes | 0.85 | 0.001 | 0.00 | 0.94 | 0.00 | 0.00 |
| LIs-MoV | 0.64 | 0.001 | 0.00 | 0.91 | 0.00 | 0.00 |
| LIs-NoC | 0.72 | 0.001 | 0.00 | 0.92 | 0.00 | 0.00 |
| LIs-NYC | 0.36 | 0.001 | 0.00 | 0.89 | 0.01 | 0.01 |
| LIs-SoT | 0.64 | 0.001 | 0.00 | 0.93 | 0.00 | 0.00 |
| LIs-Wes | 0.83 | 0.001 | 0.00 | 0.95 | 0.00 | 0.00 |
| MoV-NoC | 0.82 | 0.001 | 0.00 | 0.93 | 0.00 | 0.00 |
| MoV-NYC | 0.64 | 0.001 | 0.00 | 0.95 | 0.00 | 0.00 |
| MoV-SoT | 0.77 | 0.001 | 0.00 | 0.91 | 0.00 | 0.00 |
| MoV-Wes | 0.83 | 0.001 | 0.00 | 0.94 | 0.00 | 0.00 |
| NoC-SoT | 0.77 | 0.001 | 0.00 | 0.92 | 0.00 | 0.00 |
| NoC-Wes | 0.88 | 0.001 | 0.00 | 0.92 | 0.00 | 0.00 |
| NYC-NoC | 0.73 | 0.001 | 0.00 | 0.97 | 0.00 | 0.00 |
| NYC-SoT | 0.65 | 0.001 | 0.00 | 0.97 | 0.00 | 0.00 |
| NYC-Wes | 0.82 | 0.001 | 0.00 | 0.98 | 0.00 | 0.00 |
| SoT-Wes | 0.82 | 0.001 | 0.00 | 0.89 | 0.00 | 0.00 |

**Figure 5.**
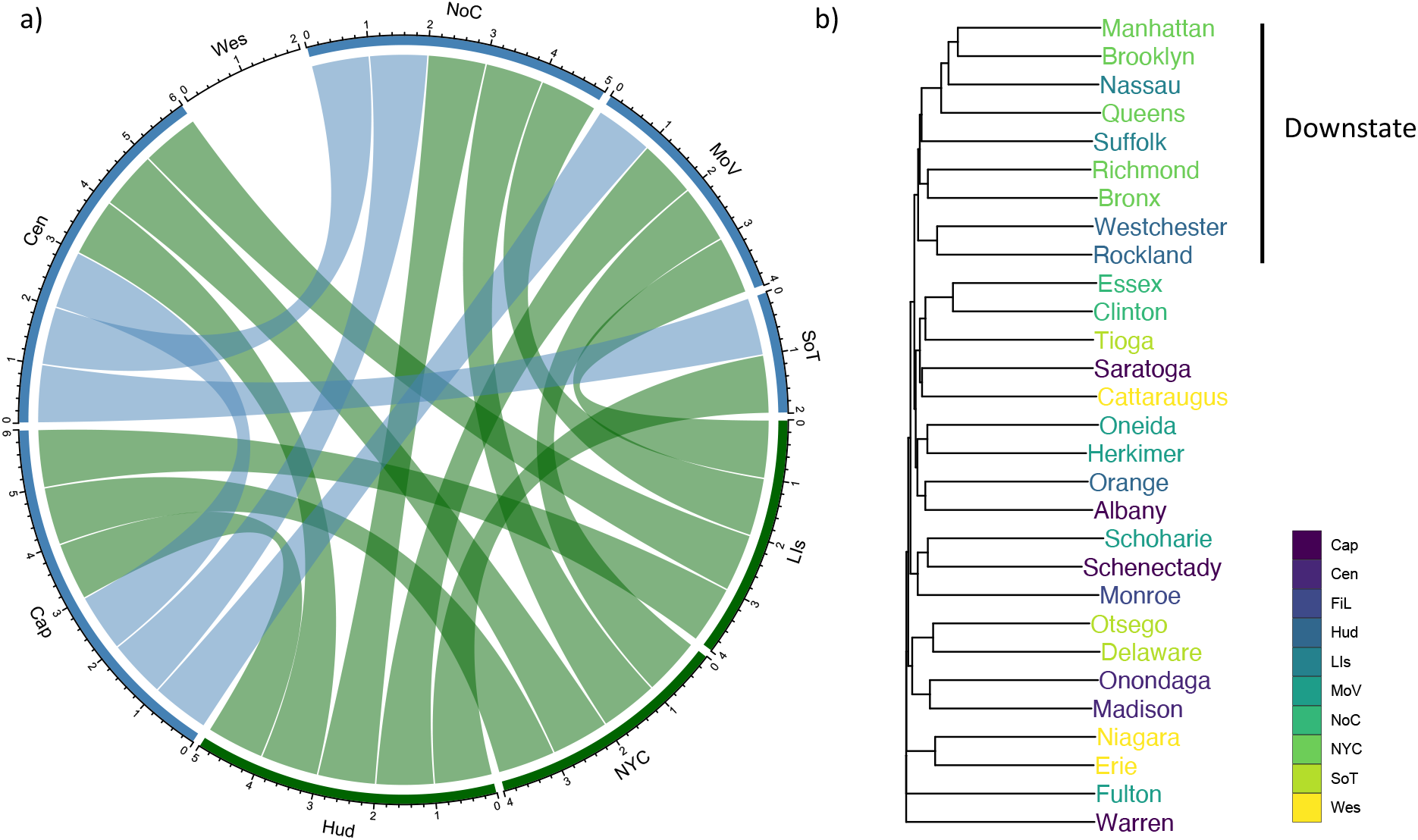
Connections among regions and counties of New York State based on SARS-CoV-2 diversity metrics. a) Circle plot of potential connections among nine of ten New York State regions as evaluated by beta-diversity metrics for the month of March. Regions that did not have significantly different SARS-CoV-2 communities (as determined by unweighted UniFrac tests), are connected by a line. Green lines are connections between downstate regions and those of upstate. Blue lines signify connections between upstate regions. While all downstate regions have at least one connection to upstate regions (except Western New York), the SARS-CoV-2 communities of many upstate regions do not appear similar. b) Dendrogram generated from Jaccard distances of shared transmission clusters among New York counties. Only counties with five sequences in one or more transmission cluster were considered. The dendrogram highlights the cohesive group formed by downstate counties. Several counties from upstate group by region but there are also outliers, suggesting transmission chains are less extensive in these areas than in the interconnected New York City metropolitan area (The five boroughs of New York City and surrounding counties). Cap, Capital District; Cen, Central; Lis, Long Island; Hud, Hudson; MoV, Mohawk Valley; NYC, New York City; NoC, North Country; SoT, Southern Tier; Wes, Western.

Examining the number of shared transmission clusters between each county/borough with more than five sequences in at least one cluster indicated a strong association of viral communities in the downstate area, including Westchester, Rockland, Nassau, Suffolk, Queens, Manhattan, Richmond, and Brooklyn (Figure 5). These results reflect the rapid spread of SARS-CoV-2 through a heavily interconnected area but contrast UniFrac analyses, which showed the viral communities of New York City, Mid-Hudson, and Long Island as significantly different from each other. The discrepancy highlights the impact of including sequences without a cluster association, which helped to distinguish downstate regions in UniFrac analyses. Counties within other regions also tended to group together based on Jaccard distances of shared clusters (Figure 5), supporting the overall distinct composition of regional SARS-CoV-2 viral communities by the end of September.

### Phylogeographic analyses reveal transmission dynamics characterized by within-state spread of SARS-CoV-2

Nextstrain phylogeographic analyses of more than 800 New York sequences and a subset of global sequences deposited on GISAID displayed the association of New York genomes with those from other states and countries (Supplementary Figure 3) as previously observed (Maurano et al. 2020; Gonzalez-Reiche et al. 2020). However, most New York genomes were concentrated in the 20C clade with an inferred date of introduction into Westchester County between mid to late February, depending on the Nextstrain build interrogated (eg. 100% CI, 2020-02-22 to 2020-02-23). These estimates are in general agreement with those of prior analyses (Worobey et al. 2020; Maurano et al. 2020) and overlap with the probable timing of infection and onset of symptoms of the index case (Gold & Ferré-Sadurní 2020).

Phylogeographic analyses conducted by BEAST indicated that up to 75% of the 303 B.1.X clusters derived from within-state transmission, although posterior probabilities for locations ranged from 0.70-1.0 depending on the Bayesian analysis (Figure 6). Llama assigned 65% of B.1.X genomes sequenced by the Wadsworth Center to a single monophyletic clade comprised of New York sequences. In other words, the majority of New York genomes sequenced by the Wadsworth Center remained most closely related to other New York sequences despite being placed within a global phylogeny. Given these results and the fact that 94% of SARS-CoV-2 genomes in New York State were of B.1 ancestry, the epidemic appeared to be fueled largely by within-state viral transmission after various initial introductions. This conclusion also agrees with epidemiological investigations of early SARS-CoV-2 cases in New York State, which found only 30% to be potentially associated with international or out-of-state travel (Rosenberg et al. 2020a), again demonstrating the effectiveness of on-pause measures limiting travel.

**Figure 6.**
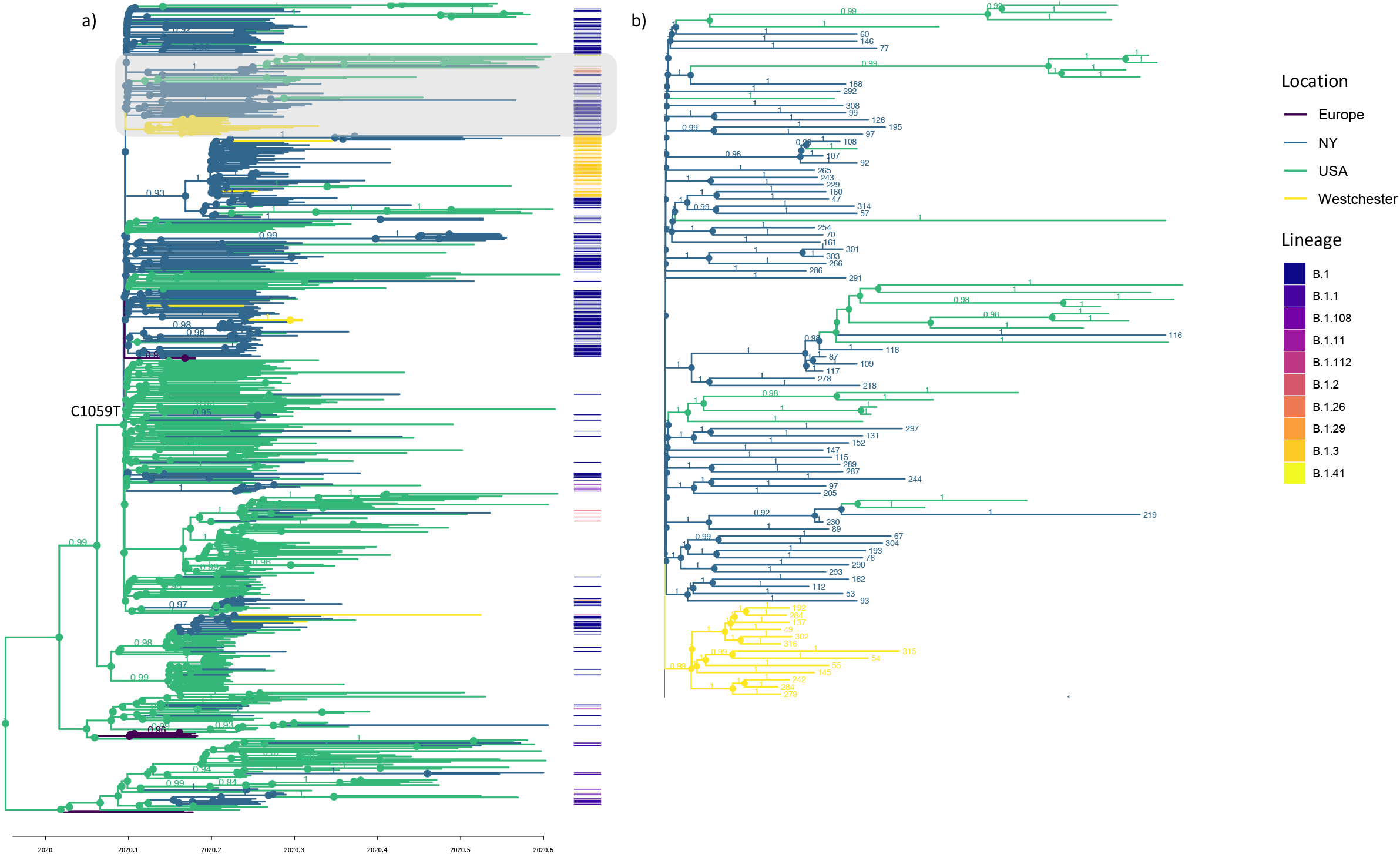
Maximum Clade Credibility tree of B.1.X SARS-CoV-2 entry in New York State. Bayesian time-calibrated phylogeny of SARS-CoV-2 genomes. New York state sequences represent a subset of transmission clusters (n=258) belonging to B.1.X Pangolin lineages sampled through the end of July. B.1.X SARS-CoV-2 genomes from the United States and Europe were randomly sampled from the same time period (with preference given for the United States). a) MCC tree with branches and nodes colored by inferred ancestral location. Associated heatmap depicts the Pangolin lineage assignments for New York SARS-CoV-2 genomes. The clade representing the Westchester outbreak is shaded in gray with genomes from the index case and epidemiologically linked genomes from Westchester in yellow. Noted is the C1059T mutation defining the 20C clade. Posterior probabilities > 0.9 are shown. b) Enlargement of the Westchester outbreak clade with tips labeled by their threshold-free transmission cluster assignment (for New York SARS CoV-2 genomes) and nodes colored by ancestral location. Ancestral location probabilities > 0.9 are shown.

A large proportion of B.1.X transmission clusters (27-50%) in the three BEAST analyses were associated with in-state viral transmission related to the Westchester County outbreak given the phylogenetic placement of the index and epidemiologically linked samples (Figure 6 and Supplementary Figure 4). The most conservative estimate derived from the third analysis containing the greatest proportion of B.1.X clusters (Figure 6). A total of 258 clusters, representing 981 genomes were represented, of which 69 clusters representing 429 genomes or 44% of all New York sequences in the analysis were related to the Westchester outbreak (not including two possible re-importation events). At a minimum, this would indicate that approximately 23% of all genomes in the entire New York dataset (429/1859) were linked to the Westchester outbreak. There was a median Hamming distance of one substitution between members of Westchester-affiliated clusters (n=429) and a median distance of four substitutions between these genomes and the rest of the dataset (n=981). We then calculated Hamming distances for 100 random samples of B.1.X sequences from Europe, the United States, and the Westchester outbreak (n=200 genomes) between March and May. The median Hamming distance between Westchester-related genomes and global representatives was also four (Supplementary Figure 5). These distances are fairly consistent with previous analyses, which found a median Hamming distance of 2 between epidemiologically linked samples and five substitutions between all globally sampled genomes (n=12814) through May with a D614G mutation (Dearlove et al. 2020).

The most conservative Bayesian estimate for the number of cases attributable to the Westchester outbreak falls between those of the threshold-free and single-linkage clustering algorithms (13.5-47%). The least conservative estimates (Supplementary Figure 4) are similar to the proportion of genomes associated with a superspreading event in Massachusetts (Lemieux et al. 2020). Even less conservatively, the Westchester outbreak would be responsible for over >76% of New York SARS-CoV-2 infections if we consider the founding of the 20C clade in the state to be a single event synonymous with the Westchester outbreak based on the defining C1059T mutation and the lack of other genomes with this signature in the United States prior to March (except one). However, our conclusions are tempered by several factors. As the number of clusters and non-New York samples increased, the number of cases associated with the Westchester outbreak decreased, highlighting the sensitivity of phylogeographic analyses to sampling strategy, particularly in the face of limited genetic diversity (Lemey et al. 2020; Frost et al. 2015). While the number of SARS-CoV-2 introductions and the clusters associated with importation events were generally consistent among the three analyses, fluctuations were observed. For example, the ancestral locations associated with B.1.2 transmission clusters changed from New York to America/USA as more closely related sequences and earlier sampled cases from other states were added (Supplementary Figure 4). Further, the phylogenetic positions of key genomes might be relatively uninformative as the small internal branch lengths and subsequent low posterior probabilities for splits within the 20C clade essentially transform it into a star phylogeny. While this could be interpreted as the rapid expansion of the Westchester outbreak it might also reflect the rapid global dissemination of the B.1 lineage, which led to transmission rates that outpaced the viral substitution rate (Bedford et al. 2020). Given the low overall diversity of SARS-CoV-2 (Dearlove et al 2020), that thousands of people might have been infected with the virus by the time of the first documented case of community transmission (Perkins et al. 2020) and that millions were estimated to be infected by the end of March (Rosenberg et al. 2020b), it is possible that closely related/identical but unsampled genomes seeded transmission chains that we ascribe to the Westchester outbreak.

All BEAST analyses provided evidence for multiple introductions of B.1.2, B.1.5, and B.1.1 lineages (Figure 6 and Supplementary Figure 4). The B.1.1 lineage was prevalent in Europe (https://cov-lineages.org/lineages.html) and our phylogeographic analyses suggest that some B.1.1 transmission chains were due to travel between New York and European countries while others originated via inter-state routes (Supplementary Figure 4). The independent introduction of the B.1.1 lineage into different New York regions supports the results from community structure analyses, which suggested that SARS-CoV-2 transmission dynamics in Western New York – predominantly influenced by the spread of the B.1.1 lineage - were largely independent of the rest of New York State.

### Conclusions

We have deposited >600 sequences from 56 out of 62 counties and all ten regions of New York to GISAID to provide a more detailed picture of statewide transmission of SARS-CoV-2 from March to September. The analysis shows the widespread distribution of the B.1 lineage and its increase in relative abundance through time, recapitulating the global trend (Korber et al. 2020; van Dorp et al. 2020). Both community structure and phylogeographic analyses suggest that a large proportion of B.1.X transmission clusters were due to within-state transmission and were related to the initial Westchester outbreak in March. From there, SARS-CoV-2 was transmitted to other downstate regions (New York City, Long Island, and other Mid-Hudson counties) and various counties of upstate New York. Indeed, the only counties that showed little relation to the initial outbreak were in the Western region of the state, but there are certainly many unsampled transmission chains. Results from phylogeographic and community structure analyses also showed downstate as a potential transmission hub. The index case associated with the Westchester outbreak had attended large religious gatherings before being confirmed as SARS-CoV-2 positive. Although around 1,000 people affiliated with the index patient were asked to self-quarantine (Jones & Maxouris 2020) and additional measures were implemented thereafter, the geographic and temporal breadth as well as the high membership of the Westchester outbreak – defined by threshold-free, single-linkage or phylogeographic strategies - underscore the consequences of even small delays in containment efforts and the disproportionate contribution large congregate settings can have in spreading the virus. However, the limited divergence of the virus combined with rapid transmission rates and under-sampling may obscure underlying sources of transmission ascribed to a single outbreak event (Worobey et al. 2020). While quantifying the number of cases related to the Westchester outbreak may not be truly possible, our application of multiple strategies to assess its impact demonstrates the importance of rapidly implementing containment measures.

Some upstate regions were dominated by lineages other than B.1 – such as Western New York, North Country, and the Mohawk Valley - frequently representing importation events and suggesting that these regions were less influenced by viral transmission from other New York areas. However, all regional SARS-CoV-2 communities became increasingly distinct since the start of the New York epidemic in March, possibly reflecting the effects of limited travel due to national restrictions and New York State on-pause measures. Thus, these data demonstrate that adherence to social distancing, mask-wearing, and the closure of schools and non-essential businesses as part of the on-pause measures were ultimately successful in curbing transmission across regions.

## Supporting information

Supplementary Tables and Acknowledgements

## Data Availability

SARS-CoV-2 genomes sequenced in this study have been deposited in GISAID.

## Acknowledgements

Next generation sequencing was performed by the Advanced Genomic Technologies Cluster of the Wadsworth Center. Funding was generously provided by the New York Community Trust. We graciously thank all originating and submitting laboratories for their SARS-CoV-2 sequence contributions to the GISAID database. We also gratefully acknowledge Dr. Joel Wertheim and Dr. Tetyana Vasylyeva for their invaluable expertise in phylodynamic analyses, Maruika Desai, MS and Michelle Hulke, PhD at the MMRI for their help in cataloguing positive specimens, and the Mohawk Valley Health System Laboratory Services for their support in sample procurement.

## IRB Approval

This work was approved by the New York State Department of Health Institutional Review Board, under study numbers 02-054 and 07-022.

## Conflicts of Interest

All authors declare no conflicts of interest.

---

**Supplementary Figure 1.**
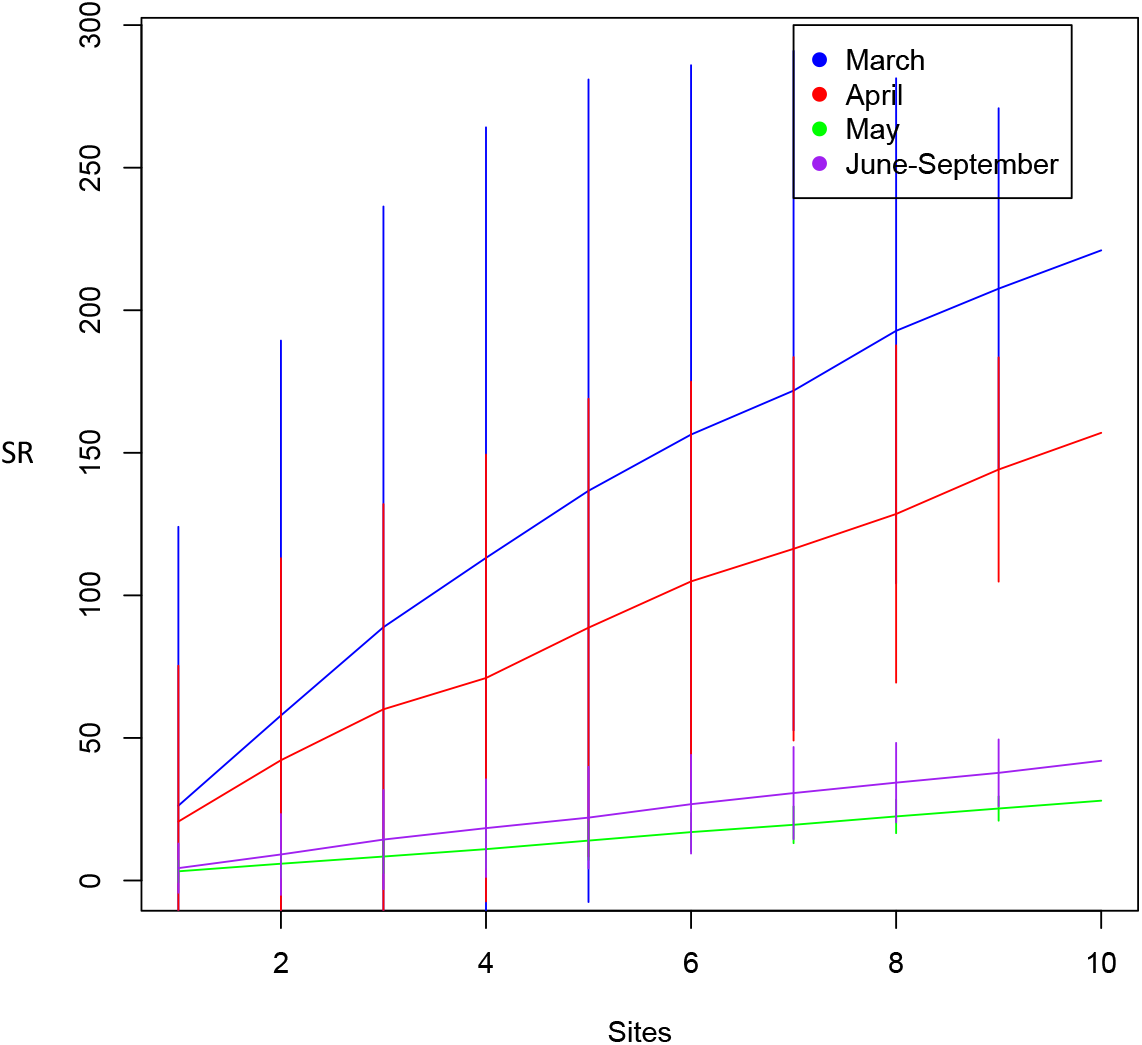
Rarefaction curves for the number of lineages detected by month as sites are randomly added to the sampling pool. Species richness (SR) of SARS-CoV-2 lineages identified as a function of the number of sites (New York State regions) sampled and the month of sampling to take into account different sampling intensities. While the number of lineages detected continues to increase as sites continue to be sampled for March and April, the curves are essentially flat for May and June-September. The rates of increase are also higher for earlier months. Vertical lines indicate standard errors associated with random permutations. Calculations were performed by Picante in R with the speaccum.psr function.

**Supplementary Figure 2.**
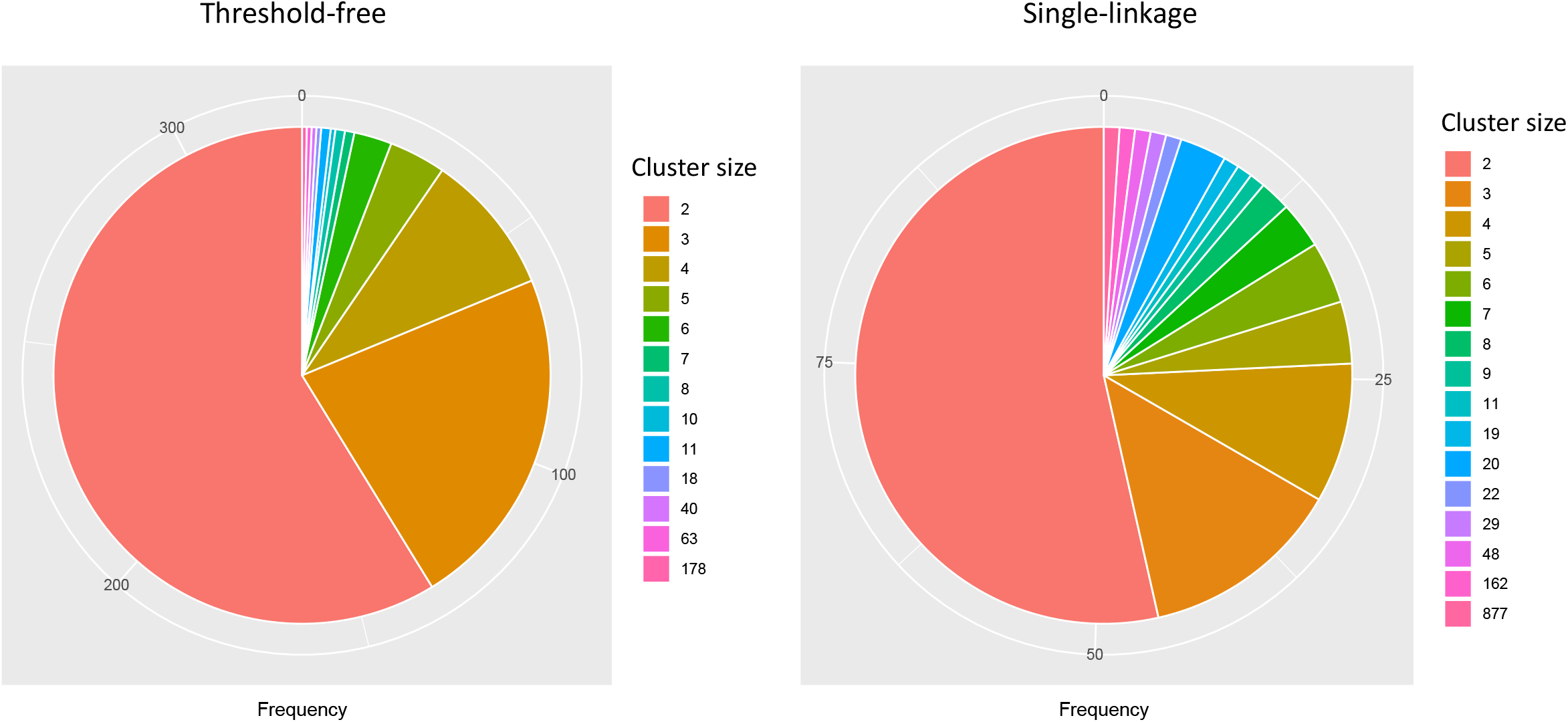
Pie graph of cluster size frequency. The number of New York State SARS-CoV-2 transmission clusters detected of different sizes by the threshold-free and single-linkage strategies.

**Supplementary Figure 3.**
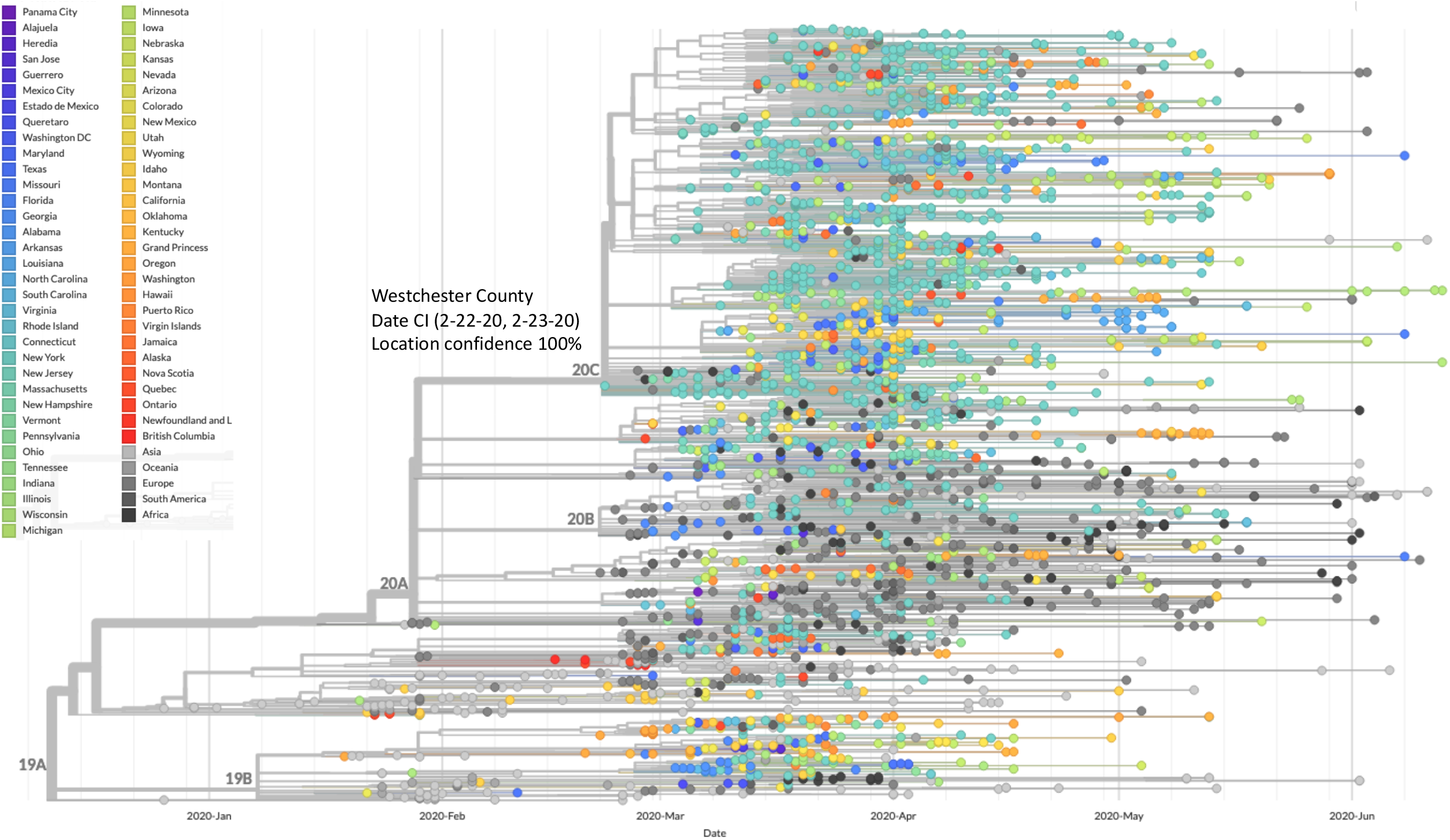
Nextstrain phylogeographic analysis of SARS-CoV-2 genomes. Representative time-calibrated phylogeny depicting evolutionary relationships among globally sampled SARS-CoV-2 genomes and those from New York State. New York state sequences are concentrated in the 20C clade and are shaded light blue.

**Supplementary Figure 4.**
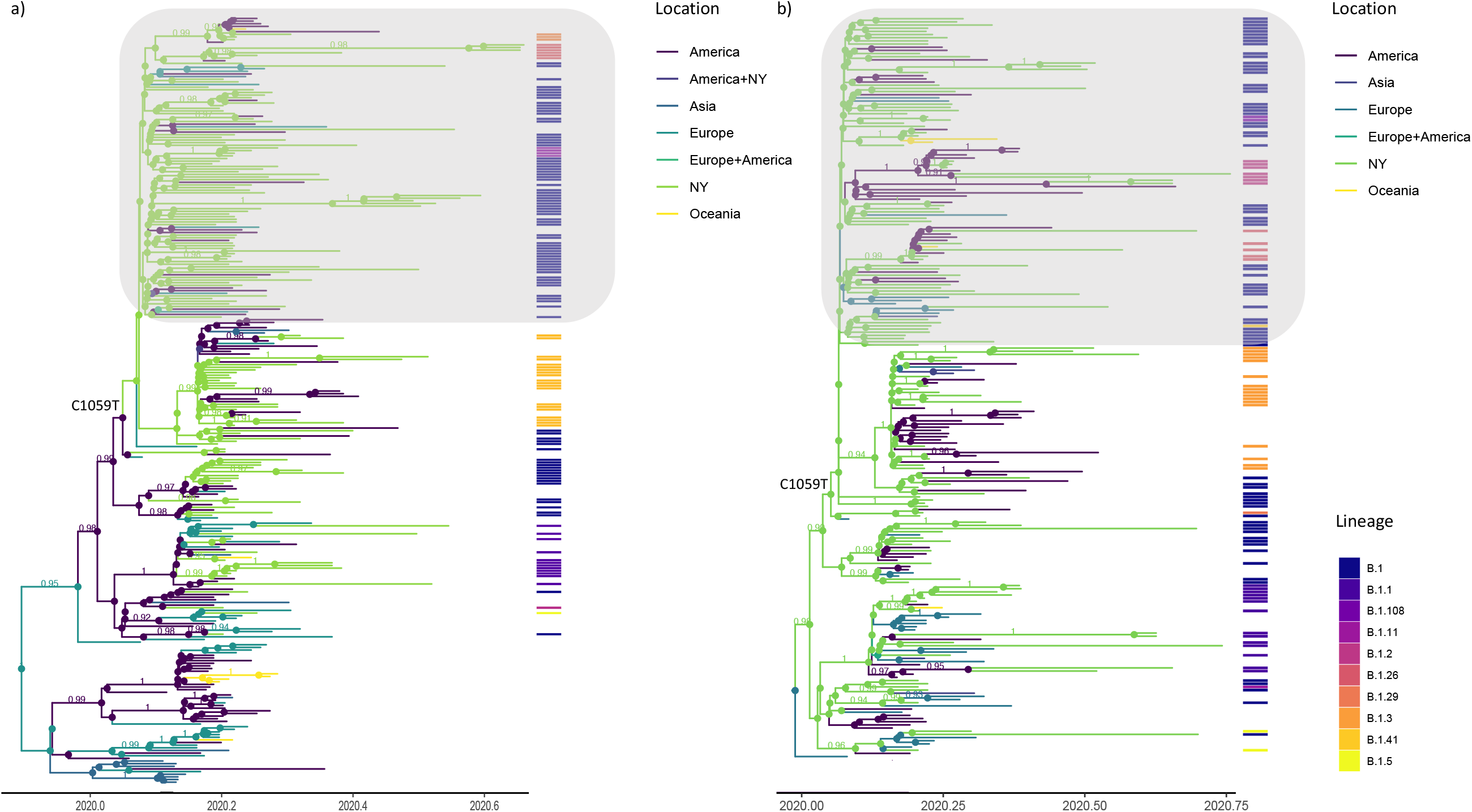
Additional Bayesian phylogeographic analyses of the B.1 lineage. Maximum clade credibility (MCC) trees including representatives from subsets of New York State B.1.X transmission clusters (a) n=94; b) n=116) and global SARS-CoV-2 genomes. Branches and nodes are colored by inferred ancestral location and posterior probabilities > 0.9 are shown. Associated heatmap depicts the Pangolin lineage assignments for New York SARS-CoV-2 genomes. The trees highlight that most B.1.X lineage clusters derive from within-state transmission and many are linked to the initial Westchester outbreak (shaded in gray) rooted by epidemiologically linked genomes to the index case. The C1059T mutation represents the defining mutation of the 20C clade.

**Supplementary Figure 5.**
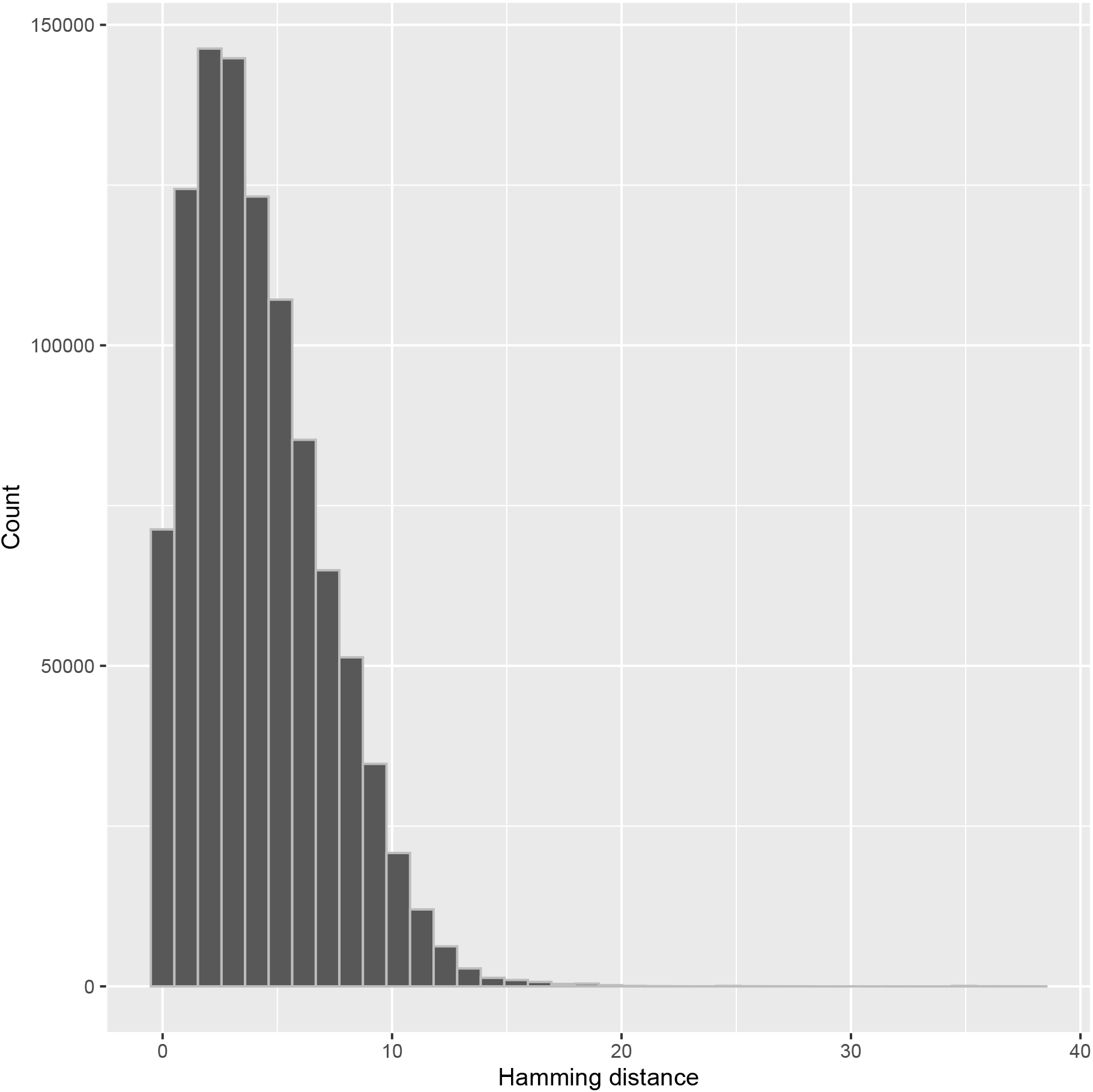
Hamming distances between SARS-CoV-2 genomes. Histogram of Hamming distances between a random subsampling of New York State genomes affiliated with the Westchester County outbreak (as defined by BEAST) and a random sampling of SARS-CoV-2 genomes from the United States and Europe through May (n=200). Calculations were performed for 100 randomizations of both datasets. The median distance between Westchester related sequences was one while the median distance between Westchester sequences and others was four.

## Notes

### Competing Interest Statement

The authors have declared no competing interest.

